# Functional Comparison of Different Exome Capture-based Methods for Transcriptomic Profiling of Formalin-Fixed Paraffin-Embedded Tumor Samples

**DOI:** 10.1101/2021.01.24.21250395

**Authors:** Kyrillus S. Shohdy, Rohan Bareja, Michael Sigouros, David C. Wilkes, Princesca Dorsaint, Jyothi Manohar, Daniel Bockelman, Jenny Z. Xiang, Rob Kim, Juan Miguel Mosquera, Olivier Elemento, Andrea Sboner, Alicia Alonso, Bishoy M. Faltas

## Abstract

**Background:** The need for fresh frozen (FF) tissue limits implementing RNA sequencing (RNA-seq) in the clinic. The majority of clinical samples are processed in clinical laboratories and stored as formalin-fixed, paraffin-embedded (FFPE) tissues. Exome capture has recently emerged as a promising approach for RNA-seq from FFPE samples. Multiple exome capture platforms are now available. However, their performances have not been systematically compared.

**Methods:** Transcriptomic analysis of 32 FFPE tumor samples from 11 patients was performed using three exome capture-based methods: Agilent SureSelect V6, TWIST NGS Exome, and IDT XGen Exome Research Panel. We compared these methods to TruSeq RNA-seq of fresh frozen (FF-TruSeq) tumor samples from the same patients. We assessed the recovery of clinically relevant biological features, including the expression of key immune genes, expression outliers often associated with actionable genes, gene expression-based subtypes, and fusions using each of these capture methods.

**Results:** The Spearman’s correlation coefficients between global expression profiles of the three capture-based methods and matched FF tumor samples, analyzed using TruSeq RNA-seq, were high (rho = 0.72-0.9, *p* < 0.05). There was a significant correlation between the expression of key immune genes between individual capture-based methods and FF-TruSeq (rho = 0.76-0.88, *p* < 0.05). All three exome capture-based methods reliably detected the outlier expression of actionable genes, including *ERBB2, MET, NTRK1*, and *PPARG*, initially detected in FF-TruSeq. In urothelial cancer samples, the Agilent assay was associated with the highest molecular subtyping agreement with FF-TruSeq (Cohen’s k = 0.7, *p* < 0.01). Both Agilent and IDT detected all the clinically relevant fusions which were initially identified in FF-TruSeq.

**Conclusion:** All exome capture-based methods had comparable performance and concordance with FF-TruSeq. These findings provide a path for the transcriptomic profiling of vast numbers of FFPE currently stored in biobanks. For specific applications such as fusion detection and gene expression-based subtyping, some methods performed better. By enabling the interrogation of FFPE tumor samples, our findings open the door for implementing RNA-seq in the clinic to guide precision oncology approaches.

## Introduction

RNA sequencing (RNA-seq) has provided deep insight into gene expression patterns in biological samples, including transcript abundance levels, isoforms expression, alternative splicing, and chimeric transcripts resulting from gene fusions. There is growing interest in leveraging RNA-seq as a clinical tool, especially in oncology, to match patients to targeted therapy and improve outcomes (1–5). One of the barriers to the clinical implementation of RNA-seq is the need for fresh-frozen tumor samples to obtain optimal results. However, in the clinical setting, the vast majority of specimens are preserved as formalin-fixed, paraffin-embedded (FFPE) tissues, which are suited for long-term storage. Unfortunately, this preservation process is associated with a rapid decline in RNA quality (2). Several adverse factors impact the quality of RNA extracted from FFPE, including ischemia, formaldehyde fixation, and embedding in warm paraffin, and the duration of the storage of tissue blocks (6,7).

RNA capture is a technique potentially more suited to the transcriptomic analysis of FFPE tumor samples (8). Recently, several commercially available RNA capture kits have become available. However, a systemic comparison of their ability to recover clinically-relevant biological features from real-world FFPE samples has not been performed. Consequently, the lack of an optimal method for transcriptomic profiling of FFPE tumor samples hinders the clinical applications. To address this knowledge gap, we compared the sequencing metrics and biological readouts from the Agilent SureSelect V6 (Agilent), TWIST NGS Exome (TWIST), and IDT XGen Exome Research Panel (IDT) capture-based methods from FFPE tumor samples. For each sample, we compared the three FFPE capture-based methods to TruSeq RNA-seq of the fresh frozen (FF) sample from the same tumor (hereafter referred to as FF-TruSeq). We studied the potential clinical utility of FFPE capture-based methods to discover clinically-useful biological readouts. The comparison focused on the outlier expression of genes, expression of key immune genes, molecular subtype classification, and gene fusions.

## Methods

### Sample collection

Patients signed informed consent (Weill Cornell IRB #1305013903). Banked excess tissue was collected from surgical specimens of patients with a diagnosis of cancer. All pathology specimens were reviewed by board-certified genitourinary pathologists in the Department of Pathology at WCM/NYP (J.M.M). Clinical charts were reviewed by the authors (K.S.S, J.M, B.M.F.) to record patient demographics, treatment history, anatomical site, and stage using the tumor, node, metastasis (TNM) system published in the AJCC Cancer Staging Manual (8th edition).

### RNA extraction methods

For RNA extraction from FFPE tissues, the Maxwell 16 ® instrument with the Maxwell® 16 LEV RNA FFPE Purification Kit was used as previously described (9). This kit provides a high yield of pure RNA from FFPE tissue (and FF tissue, see below) samples. This protocol takes 60 minutes after macrodissection of the unstained FFPE slides and Proteinase K digestion to complete. Prior to macrodissection, hematoxylin and eosin (H&E) stained slides were cut and annotated by a pathologist to identify the location of the tumor in the corresponding unstained slides to be used in the extraction. Ten unstained slides of 10 µm thickness per case were cut for the extraction along with one H&E stained slide. The annotated locations on each slide were then macro-dissected with a sterile razor blade to obtain tissue for RNA extraction.

For extraction from frozen tissue, the Maxwell 16 ® instrument with the Maxwell® 16 LEV simplyRNA Tissue Kit was also used. Similarly, H&E stained slides were cut from the corresponding frozen block and annotated by a pathologist to identify the tumor location. Tissue from these annotated locations was removed using 1.5 mm diameter punch biopsies to core the block. Tissue homogenization was aided by introducing stainless steel beads to the tissue/homogenization solution and using the Qiagen Tissue Lyser LT set at 1/50s for 2 minutes to physically break up the tissue before the lysis buffer was added.

### RNA quantity and quality assessment

The quantity of RNA was determined using a Nanodrop 2000 for nucleic acid absorbance measurement and a Qubit Fluorometer to confirm RNA concentration (ThermoFisher, Waltham, MA). Quality was assessed using a Bioanalyzer2100 (Agilent Technologies, Santa Clara, CA) with a high sensitivity RNA chip. The RIN number was used to decide which RNA library prep kit to use for the frozen tissues; the DV_200_ measurement (the % of RNA fragments >200nt) was used to determine the degree of RNA fragmentation for the FFPE samples (Evaluating RNA Quality from FFPE Samples. Illumina, Technical Note, publication number 470-2014 001. https://www.illumina.com/content/dam/illumina-marketing/documents/products/technotes/evaluating-rna-quality-from-ffpe-samples-technical-note-470-2014-001.pdf); SureSelectXT RNA Direct Protocol Provides Simultaneous Transcriptome Enrichment and Ribosomal Depletion of FFPE RNA, Agilent Technologies, Technical Note, publication number PR7000-0679. (https://www.agilent.com/cs/library/applications/5991-8119EN.pdf)

### To define the impact of the quality of the extracted RNA from FFPE samples on uniquely mapped reads

The relationship between two critical quality metrics was analyzed, the percentage of fragments >200 nucleotides (DV200 values) and RNA Integrity Number (RIN). For all FFPE tumor samples, DV200 and RIN ranged from 22-87 (median 45) and 2-2.7 (median 2.4), respectively (**Supplementary Table 1 and 2, and Supplementary Fig. 1a, b**). Across the same patient tumor samples, the RIN values were significantly correlated with DV200 (Spearman’s r = 0.54, *p* = 0.0015) (**Supplementary Fig. 1c**). Samples with DV200 of <20, 20-30, or >30 had a similar degree of correlation with the number of uniquely mapped reads and the mapped reads percent. Overall, the DV200 showed no significant correlation with the number of mapped reads (Spearman’s r = 0.13, and 0.16, *p* = 0.51 and 0.37, respectively) **(Supplementary Fig. 2a, b)**, suggesting that low DV200 does not significantly impact the sequencing metrics of the FFPE capture-based methods. Similarly, the RIN value of each FFPE tumor sample did not lead to a significant difference among the uniquely mapped reads or the mapped reads percentage from the three FFPE capture-based methods (Spearman’s r = 0.12, and 0.12, *p* = 0.45 and 0.22, respectively) (**Supplementary Fig. 2 c, d**). Both RIN and DV200 had a limited utility for excluding low-quality samples for exome capture-based methods.

### Construction of RNA-seq library and sequencing

#### RNA library preparation from fresh frozen tumor tissues

For RNA with RIN≥6.0, libraries were prepared using TruSeq RNA Library Prep kit v2 (Illumina, San Diego, CA, PN-RS-122-2001). Briefly, poly A+ RNA was purified from 100 ng of total RNA with oligo-dT beads and fragmented to ∼200bp. cDNA was synthesized using random priming, then end-repair, dA-tailed, and ligated to Illumina TruSeq adaptors containing unique sequencing indexes. Libraries were amplified with 15 cycles of PCR and pooled for sequencing (**Supplementary Tables 1 and 2)**.

For RNA with RIN<6, libraries were prepared with TruSeq Stranded Total RNA kit (Illumina, San Diego, CA, PN-20020596). Briefly, rRNA was depleted from 200ng of total RNA with Ribo-Zero and fragmented to ∼200bp. cDNA was synthesized using random priming, and transcript orientation was preserved by using dUTP during second-strand cDNA synthesis. After end repair, A-tail, and ligation to Truseq adapters, libraries were generated by amplification with 15 cycles of PCR.

Library pools were clustered at 6.5pM on a paired-end read flow cell and sequenced for 75 cycles on an Illumina HiSeq 2500 to obtain ∼50M reads per sample. **(Supplementary Table 1 and 2)**.

### RNA-exome capture libraries

Briefly, stranded RNAseq libraries were generated as per the manufacturer’s recommendations, but without the transcriptome enrichment step (pre-capture libraries). Transcriptome enrichment was achieved by the hybridization of the pre-capture library to the exome panels tested. Since the probe baits were biotinylated, hybridized libraries were captured using streptavidin beads (ThermoFisher, Waltham, MA) and PCR amplified-on-beads to generate a post-capture library. All post-capture libraries were subjected to quality control on an Agilent Bioanalyzer and normalized to 2nM. The post-capture libraries obtained from each capture platform were pooled, and each pool sequenced on one lane of a paired-end read flow cell for 2×100 cycles on a HiSeq4000 to obtain ∼40M reads per sample. The primary processing of sequencing images was done using Illumina’s Real Time Analysis software (RTA). CASAVA 1.8.2 software was then used to demultiplex samples and generate raw reads and respective quality scores (**Supplementary Tables 1 and 2)**.

### Sure Select^XT^ Human All exon v6+UTRs

(PN-5190-881, Agilent, Santa Clara, CA): Non-indexed pre-capture libraries were made using SureSelect ^XT^ RNA Direct kit (PN-G9691-90050) with 200 ng of RNA, using the % DV_200_ obtained with the Agilent Bioanalyzer to determine fragmentation times and amplifying 14-16 PCR cycles. Hybridization was carried out with 200ng from each pre-capture library for 24hrs x 65°C on RNA-biotinylated probes. Indexes were added during post-capture libraries amplification using 12 cycles.

### Twist Human Core Exome

(PN100790, Twist Biosciences, San Francisco, CA): Libraries were made using the NEBNext Ultra II Directional kit (PN-E7760, New England Biolabs, Ipswich, MA) with 100 or 200ng depending on the % DV_200_ of the starting material. Pre-capture libraries were uniquely indexed for Illumina sequencing, using 11-16 amplification cycles. 1.5µg of pooled indexed libraries (300ng each, two pools) were hybridized to the biotinylated double-stranded DNA probe panel for 16hrs at 70°C. Post-capture libraries were amplified for 8 cycles (DOC-001014).

### IDT xGen Exome Research Panel v1.0

(Integrated DNA Technologies, Coralville, IA): Libraries were made as described above using the NEBNext Ultra II Directional kit (PN-E7760, New England Biolabs, Ipswich, MA). 5µg of pooled indexed libraries (500ng each) were hybridized to the biotinylated oligo probes for 4hrs at 65oC. Post-capture libraries were amplified for 7 cycles (NGS-10122-PR 01/2020).

### RNA Sequencing Analysis

All reads were independently aligned with STAR_2.4.0f1 (10) for sequence alignment against the human genome sequence build hg19, downloaded via the UCSC genome browser [http://hgdownload.soe.ucsc.edu/goldenPath/hg19/bigZips/], and SAMTOOLS v0.1.19 (11) for sorting and indexing reads. Cufflinks (2.0.2) (12) was used to estimate the expression values (FPKMS), and GENCODE v19 (13) GTF file for annotation. Since the sequenced samples were processed using different library preps, batch normalization of FPKMs from WCM Frozen samples was done using ComBat from the sva Bioconductor package (14). For fusion analysis, we used STAR-fusion (STAR-Fusion_v0.5.1)(15,16). Fusions with significant support of junction reads (≥1) and spanning pairs (≥1) were selected. For outlier detection, the FPKMs from batch normalized frozen WCM samples were combined with the FPKMs from FFPE samples. We only selected the druggable genes from drugbank (17) as well as cancer genes from Oncokb (18), which resulted in a list of 138 druggable cancer genes. The mean and standard deviation of each gene were calculated across the WCM RNA-seq cohort (multiple cancer types). An outlier was defined as having 1.5 times the interquartile range, z-score ≥2, and FPKMs ≥20.

### Statistical analysis

For pairwise comparisons, we used the Wilcoxon signed-rank test. For comparison of the post-alignment statistics among the three capture methods, the Kruskal-Wallis test was performed. Correlation analyses between gene expression profiles were performed using Spearman’s rank correlation. For analysis of the inter-classifier agreement, Cohen’s kappa statistic measure of inter-rater agreement was calculated. The kappa-statistic measure of agreement was scaled to be 0 when the amount of agreement is what would be expected to be observed by chance and 1 when there is perfect agreement. For intermediate values, we used the Landis and Koch (19) suggest the following interpretations: below 0.0 Poor, 0.00 –0.20 Slight, 0.21 –0.40 Fair, 0.41 –0.60 Moderate, 0.61 –0.80 Substantial, 0.81 –1.00 Almost perfect. RStudio (1.0.136) with R (v3.3.2) and ggplot2 23 (2.2.1) were used for the statistical analysis and the generation of figures. A *p-value* < 0.05 was considered significant.

## Results

### Overview of the study

We conducted this study to answer two main questions. First, Among the three commercially available FFPE capture-based methods (Agilent, TWIST, IDT), what are the performance characteristics of the three methods compared to each other? Second, what are the performance characteristics of FFPE capture-based methods compared to TruSeq of their matched FF tumor samples (**Fig. 1**)?

**Figure 1.**
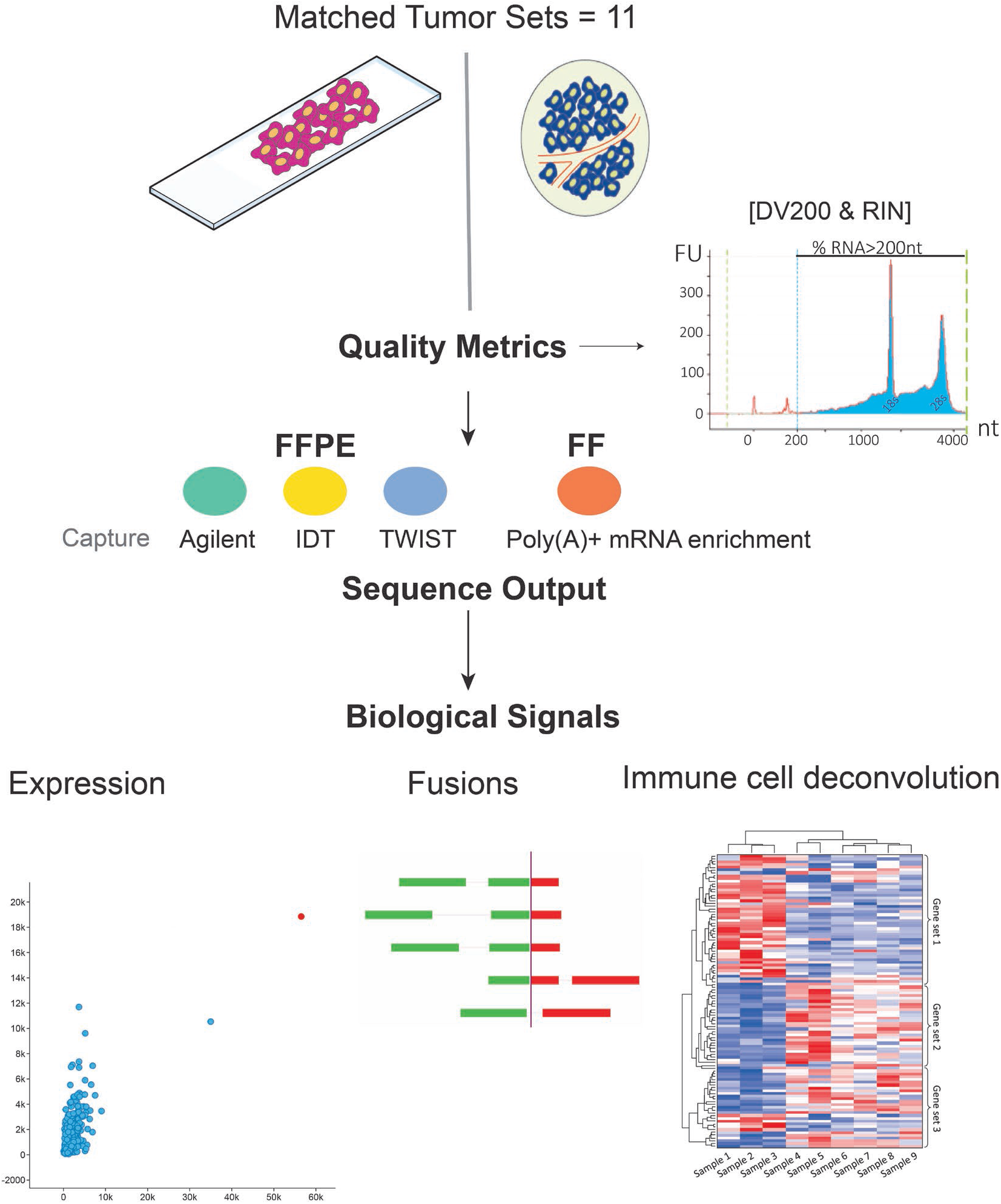
Study overview. Three exome capture-based methods (Agilent, TWIST, IDT) were used to construct sequencing libraries from FFPE tumor samples and were compared. The performance of the three capture-based methods in identifying readouts such as outlier gene expression, fusions, and immune gene expression was benchmarked against FF-TruSeq of their matched fresh frozen samples from the same respective tumor.

To answer these questions, we compiled a cohort of 32 FFPE tumor samples from 11 patients. For each patient, a matched FF tumor sample was available. We included several tumor types, namely urothelial cancer, gastroesophageal junction (GEJ) adenocarcinoma, oligodendroglioma, cancer of unknown primary (CUP), leiomyosarcoma, papillary thyroid cancer, and colorectal cancer. (**Supplementary Table 3**). We performed RNA seq (capture-based methods and TruSeq) of FFPE and FF tissues from the same tumor samples (**Fig. 1**).

The Agilent capture-based method targets 91 Mb of genomic DNA sequence in addition to 5’ and 3’ UTR sequences. IDT and TWIST methods target 39 and 33 Mb of the coding sequences (CDS) of human coding genes, respectively. The three capture-based methods use 120-base RNA probes to capture known coding DNA sequences. The total number of captured genes for the three methods are 20,456 (Agilent), 19,075 (IDT), and 19,542 (TWIST).

### Alignment statistics

To compare the performance of FFPE capture-based methods, we analyzed mapping statistics and compared them with TruSeq of matched FF tumor samples. The mean number of input reads was 38.6 million for FFPE capture-based methods and 44.4 million for FF-TruSeq. The mean number of input reads was not statistically significant between the capture-based methods **(Fig. 2a and Supplementary Table 4)**. The mean total number of uniquely mapped reads was 35 million for FFPE capture-based methods and 39 million for FF-TruSeq. The mapped reads percentage (the ratio of mapped reads to the input reads) was high for FFPE capture-based methods (mean 91.33%, SD = 3.20) (see **Methods)**. Across the FFPE capture-based methods, the mapped reads percentages were comparable between Agilent and IDT (Wilcoxon rank *p* > 0.05) and IDT and TWIST (Wilcoxon rank *p* > 0.05) (**Fig. 2b)**. TWIST was associated with a significantly lower percentage of mapped reads (89%) compared to Agilent (94%) (Wilcoxon rank *p* = 0.036) **(Fig. 2b)**. The percentage of multi-mapped reads were low across all FFPE capture-based methods (mean 3.44%, SD = 1.71). The Agilent capture method was associated with the lowest percentage of multi-mapped reads (2%) compared to IDT (5%, Wilcoxon rank *p* = 0.00013) and TWIST (3%, *p* < 0.0001) **(Supplementary Fig. 3a and Supplementary Table 4)**. Altogether the mapping metrics were comparable across capture-based methods and FF-TruSeq.

**Figure 2.**
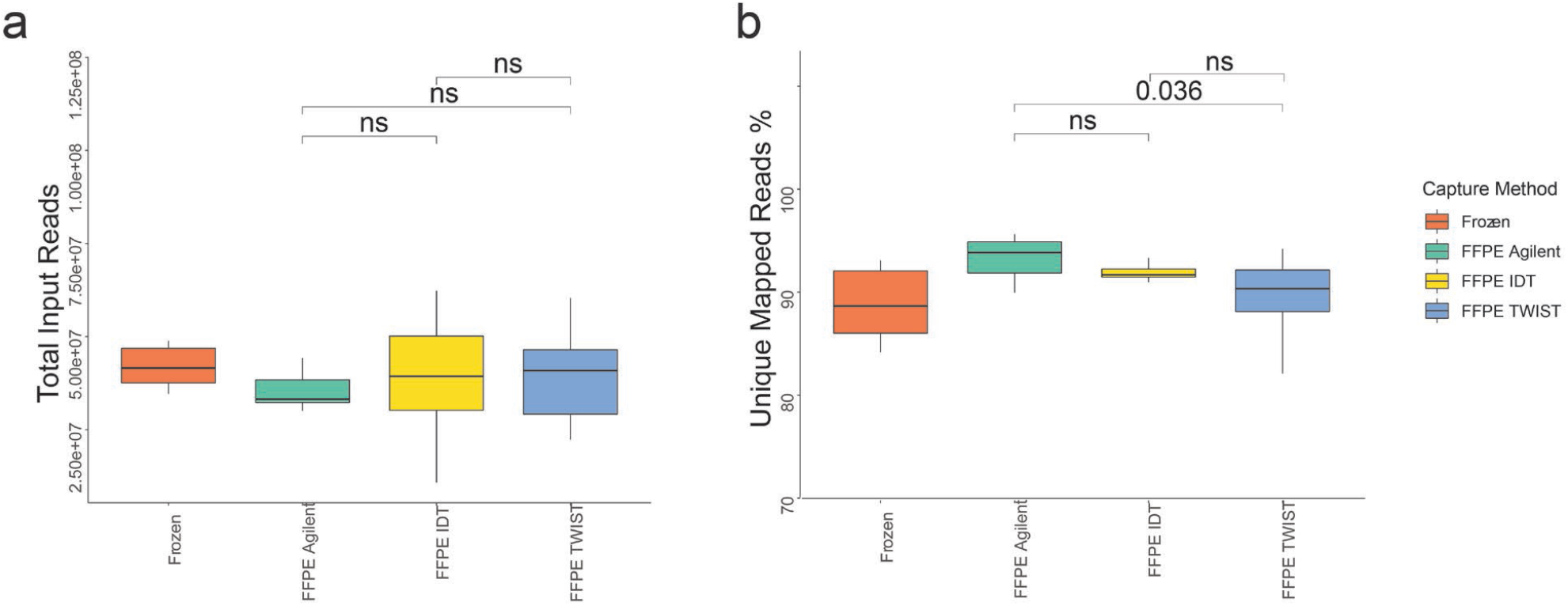
Sequencing outputs were comparable among three capture-based methods. (a) Boxplots comparing the number of total input reads show no significant difference among three exome capture-based methods. (b) Boxplots comparing uniquely Mapped reads percentage among three capture-based methods.

### Global mRNA expression

We defined the mRNA expression levels using FPKMs (fragments per kilobase of exon model per million reads mapped) from capture-based methods. TWIST showed the highest median log FPKMs compared to IDT (*p* < 0.0001) and Agilent (*p* < 0.0001) **(Supplementary Fig. 3b)**. We examined the distribution of FPKMs from capture-based methods. The mRNA gene expression from FF RNA-seq is known to follow bimodal distribution (20–22). Consistent with FF, we found that the three FFPE capture-based methods had two major density peaks, with the first density peak of genes at 0 FPKM per gene and another with 1000 FPKM per gene. Similarly, the distribution of gene expression of FF-TruSeq was bimodal, showing one peak density at 0 FPKM per gene and another peak of genes at 100 FPKM per gene **(Supplementary Fig. 3c)**. Overall, the percentage of genes with no detectable expression was not significantly different between the three capture methods and the FF-TruSeq **(Supplementary Fig. 3d)**. It is noteworthy that the three FFPE capture-based methods captured a total of 17,801 genes that were common among the three methods. The unique genes that were captured by each method were 1880 for Agilent, 360 for TWIST, and 216 for IDT (**Supplementary Fig. 3e)**.

We then asked whether the expression profiles of the FFPE capture-based methods matched the FF-TruSeq profiles derived from the same samples. All of the global expression profiles of the FFPE had a significant correlation with the corresponding FF-TrueSeq from the same tumor sample (Spearman’s r 0.72-0.90, *p* < 0.05) **(Fig. 3)**. In one patient (R11), the Agilent capture method showed a lower Spearman’s correlation of 0.72 with the corresponding FF-TruSeq-FF sample (*p* < 2.2e-16). Overall, the global gene expression pattern of FFPE tumor samples clustered with the corresponding FF sample in 8/11 of the matched sample sets in t-distributed stochastic neighbor embedding (t-SNE) plot (**Supplementary Fig. 4**). In only one patient (R11), the three FFPE capture-based methods did not cluster together **(Supplementary Fig. 4)**. These data suggest that the capture-based methods provide a gene expression profile that correlates well with FF-TruSeq.

**Figure 3.**
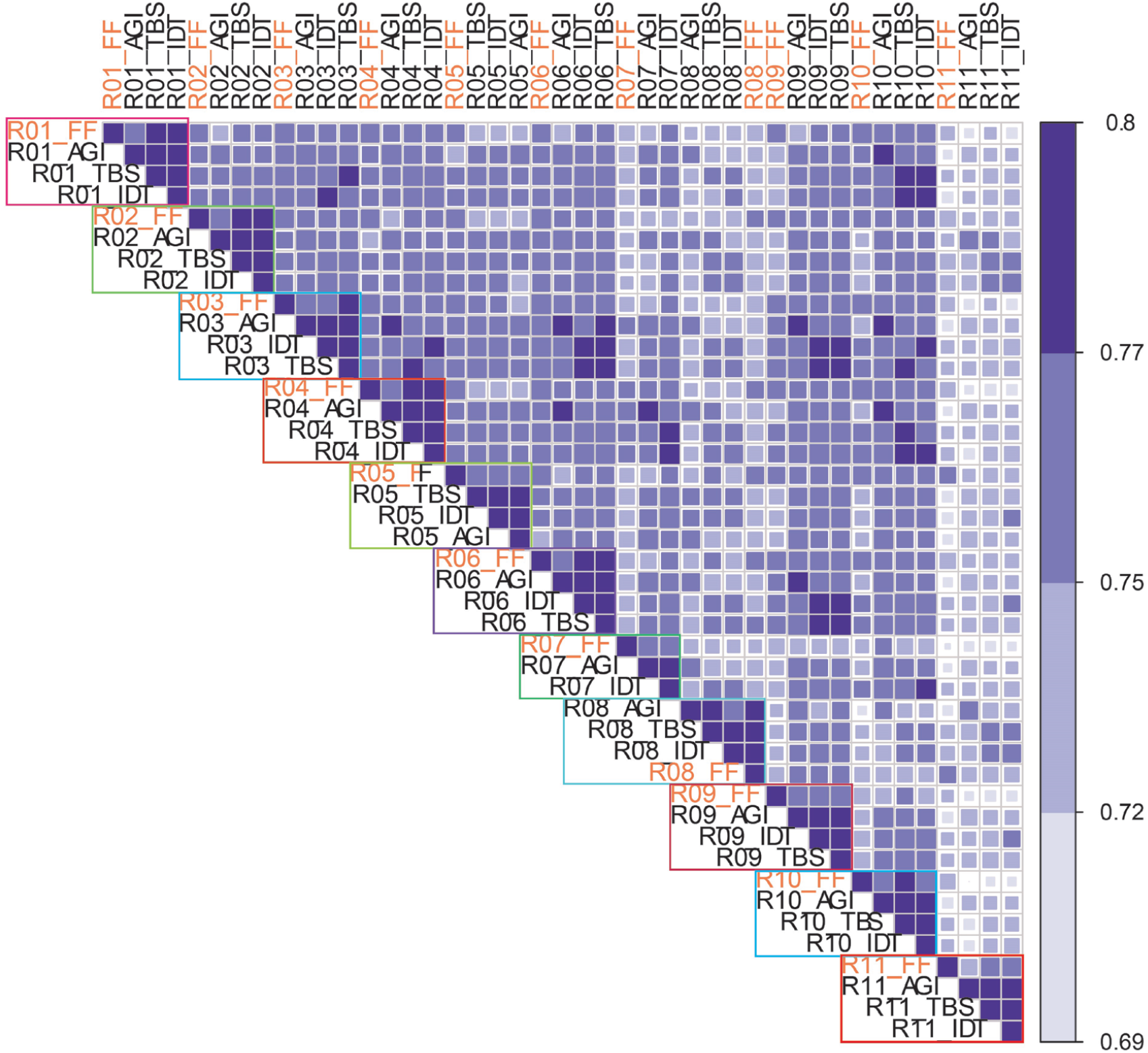
Heatmap figure showing the Spearman’s correlation coefficient of global mRNA expression. Each outlined block represents samples from the same patient. Color scale: 0.69 (lighter blue color) to 0.8 (darker blue color).

Cancer cells exhibit outlier expression of several oncogenic transcripts. These overexpressed transcripts are potential targets (23). We measured the concordance of the expression of outlier clinically relevant genes in FF-TruSeq and compared whether the same outliers could be recovered from FFPE capture-based methods. For outlier detection, the mean and standard deviation of a gene was calculated across the WCM RNA-seq cohort consisted of 650 multiple tumor samples. Outlier expression was defined as 1.5 times the interquartile range, z-score ≥ 2, and FPKM ≥ 20 (see **Methods)**. *ERBB2* was found to be an outlier in three urothelial cancer patients, including the three FFPE capture-based methods and FF-TruSeq (**Fig. 4a**). *MET, NTRK1*, and *PPARG* showed outlier expression in three patients with GEJ adenocarcinoma, colorectal cancer, and urothelial cancer, respectively. There was 100% concordance for outlier detection between FFPE capture-based methods and FF-TruSeq. These data suggest that FFPE capture-based methods provide a reliable tool to identify gene expression outliers that are clinically-relevant.

**Figure 4.**
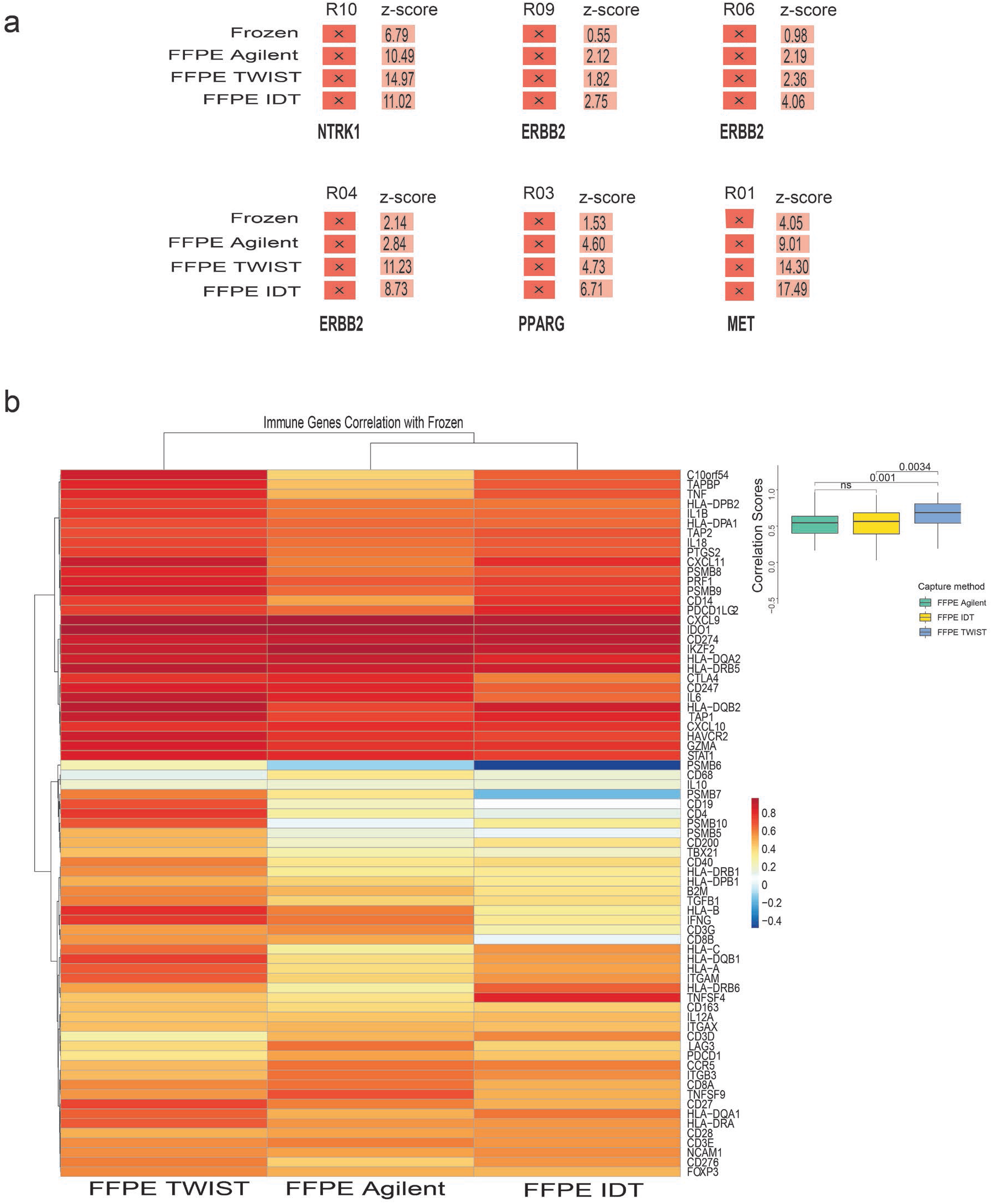
Gene expression outliers and immune gene correlation. (a) Outlier genes (and z-scores) in FF-TruSeq and the corresponding samples from the capture-based methods in the same patient. (b) Heatmap showing Spearman’s correlation coefficients based on gene expression of immune genes. Each cell shows correlation coefficients of all the samples from a capture method with corresponding FF-TruSeq. Overall, TWIST shows a higher correlation with FF-TruSeq.

### Measurement of mRNA expression of immune genes

Characterization of the immune infiltration using gene expression provide a wide scale of information in many cancer types and have prognostic and predictive values (24). For instance, the expression of immune checkpoint genes correlates with response to immune checkpoint blockade in a wide range of cancer types (25–28). We quantified the concordance of the expression of 73 key immune genes using FPKM values (see **Methods)** in FFPE exome capture-based methods and FF-TruSeq. A heatmap of the Spearman’s correlation scores among the expression profile from the three FFPE capture-based methods and FF-TruSeq is shown in **Fig. 4b**. Overall, the expression of individual genes in FFPE capture-based methods correlated with expression in matched FF-TruSeq. The expression of PD-L1 (CD274) and CTLA4 in FF-TruSeq significantly correlated with expression in Agilent (r= 0.85, *p* =0.0021 and r= 0.83, *p* = 0.0031), IDT (r = 0.87, *p* = 0.00095 and r = 0.88, *p* = 0.00067), and TWIST (r= 0.76, *p* = 0.016 and r = 0.88, *p* = 0.002), respectively **(Supplementary Fig. 5)**. Overall, TWIST was associated with the highest correlation scores with the frozen, which were significantly higher than Agilent (*p* = 0.001) and IDT (*p* = 0.0034) **(Fig. 4b)**. These results suggest that FFPE exome capture-based methods provide a practical alternative to determine the expression of immune genes from tumor samples.

### mRNA expression-based molecular classification of bladder cancer

A consensus mRNA expression-based single-sample classifier of muscle-invasive bladder cancer was recently published (29). Applying this classifier to 18 datasets, six molecular classes were previously identified: luminal papillary (LumP), luminal nonspecified (LumNS), luminal unstable (LumU), stroma-rich, basal/squamous (Ba/Sq), and neuroendocrine-like (NE-like) (29). To assess the applicability of using this classifier on RNA-seq data from FFPE tumor samples, we measured the concordance of the classifier between the three FFPE capture-based methods and FF-TruSeq.

We applied this classifier to the transcriptomic data of FF-TruSeq urothelial tumor samples and their matched FFPE capture-based methods from five patients. The three capture methods showed significant agreement with FF (50%-80%) in classifying the molecular subtype (**Supplementary Table 5**). The Cohen’s kappa for the agreement between consensus molecular classes was moderate to perfect for LumP (0.6), LumU (0.7), and Basal/squamous (1.00), whereas it was slight to poor for Stroma-rich (0.2) and LumNS (−0.1). The NE-like subtype was not represented in our dataset.

### Fusion detection

We evaluated the performance of the exome capture-based methods Agilent, TWIST, and IDT in detecting gene fusions compared to FF-TruSeq. In our cohort, we selected eight fusion transcripts that were initially identified in FF tumor samples (see **Methods)**. Four fusions (e.g., NCOA4-RET, CCDC6-RET, TPM3-NTRK1, and MKRN2-PPARG) were orthogonally confirmed with targeted sequencing using the Archer FusionPlex platform from the FF samples (30). The three FFPE capture-based methods detected all the fusions except for the MKRN2-PPARG fusion, which was not detected by the TWIST capture method in one sample **(Fig. 5a)**. In the FFPE tumor samples, the junction reads significantly correlated with the expression of the fusion transcripts (r = 0.95, *p* < 000.1). The Spearman’s correlation coefficients between junction reads and expression levels were 0.99 (Agilent, *p* < 0.0001), 0.92 (TWIST, *p* = 0.001) and 0.85 (IDT, *p* = 0.0034). respectively. The junction reads and spanning supporting each fusion across the three capture methods were comparable with FF-TruSeq **(Fig. 5b and Supplementary Table 6**). Collectively, these data indicate that the FFPE capture-based methods can identify the majority of fusions.

**Figure 5.**
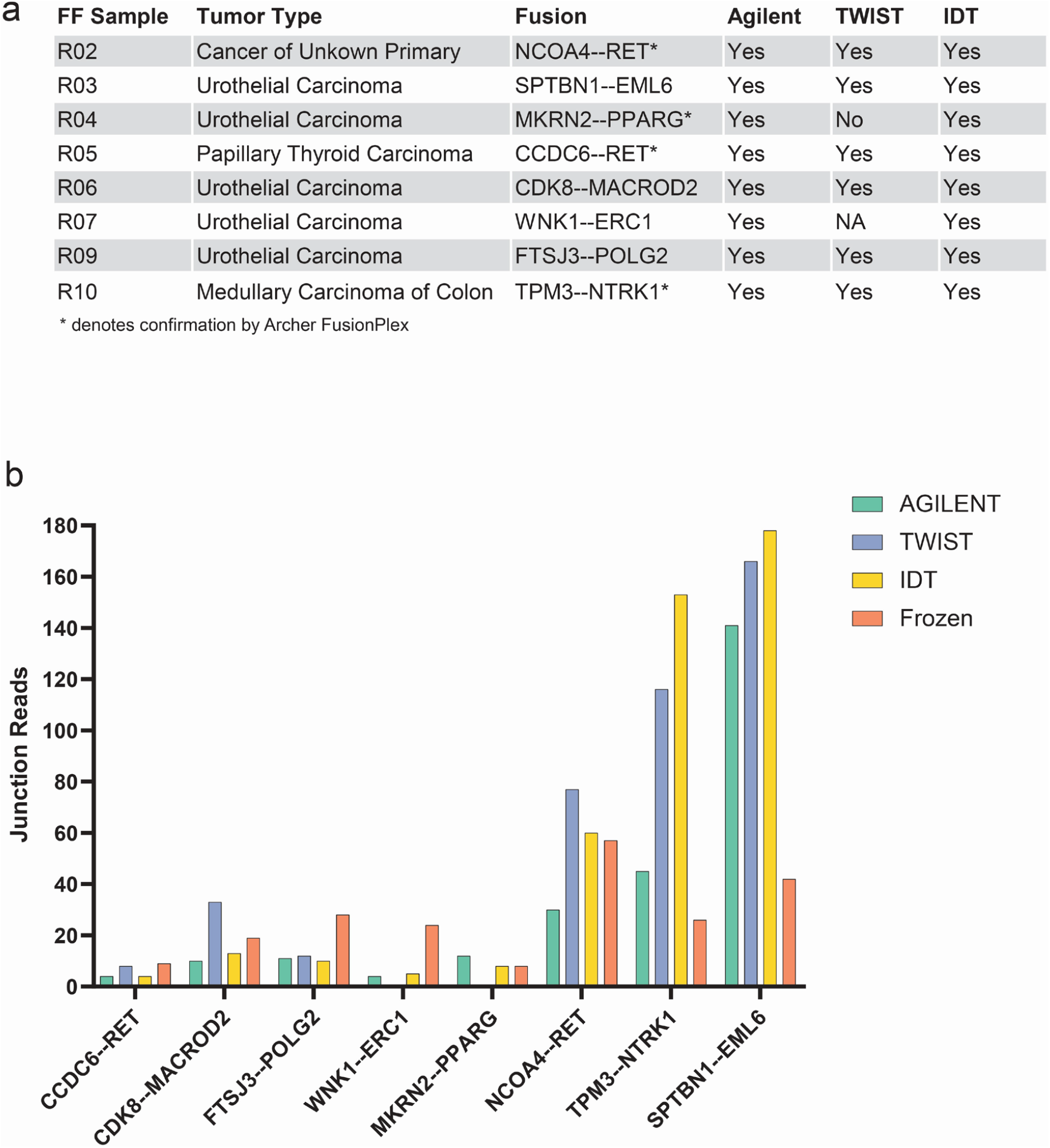
Detection of oncogenic gene fusions. a) Table showing eight gene fusions that were identified in the FF tumor samples and their detection status in the corresponding capture-based methods. No sample was available for TWIST for one patient (R07). b) Bar plots of the junction reads supporting the eight identified fusions derived from FFPE capture-based methods and matched FF-TruSeq.

## Discussion

RNA-seq has many advantages, including simultaneously measuring the expression of thousands of genes to provide functional signal pathway activation or suppression and detection of oncogenic gene fusions. For these reasons, it provides a valuable addition to the precision medicine toolkit.

In this study, we performed transcriptomic profiling of FFPE tumor samples using three capture-based methods (Agilent, TWIST, and IDT). We benchmarked the three capture-based methods with the TruSeq from matched FF tumor samples. Our diverse tumor cohort recapitulated the real-world FFPE biobanks and supported the generalizability of results to other cancer types.

The FFPE capture-based methods showed high performance in identifying biological signals, including outlier genes, oncogenic fusions, and key immune gene expressions. Other biological signals need to be interpreted cautiously, such as the molecular subtype classification for specific subtypes. Since it is based on a composite gene expression pattern, it is inherently more complicated than a single gene comparison.

The three capture-based methods successfully generated sequencing libraries for all tumor sample sets. The peak density of the DV200 and RIN were within the accepted quality range to proceed with library prep. Moreover, samples with low-quality metrics indicating highly degraded specimens did not adversely impact sequencing output or the number of uniquely mapped reads. Low DV200 and RIN are not sufficient to exclude samples from sequencing for a given capture method.

The differences in the total number of captured genes among the three capture-based methods did not change the global gene expression profile. Furthermore, the global expression was positively correlating with the FF-TruSeq sample across the 11 matched tumor sets, suggesting the reliability of the transcriptomic-based information derived from FFPE capture-based methods compared to FF-TruSeq.

We performed several downstream analyses on the transcriptomic data to show the comparable clinical utility of FFPE to FF. We focused our analysis on clinically meaningful biological readouts, including the detection of expression outliers and oncogenic gene fusions (both are amenable for therapeutic strategies) along with the expression-based molecular classification of tumors that have prognostic value (31).

Identifying targetable outlier genes from RNA-seq has important clinical applications. *ERBB2* was found among the outlier genes in three tumor sample sets from three urothelial cancer patients. One patient showed an exceptional clinical response to trastuzumab following the detection of *ERBB2* expression (32).

Two oncogenes *MET* and *NTRK1*, with FDA-approved targeted therapies (capmatinib and larotrectinib), were also identified in a patient with GEJ and another patient with medullary colon cancers, respectively. Our data suggest that gene expression from FFPE samples is a potential tool for patient selection for oncogene-targeted therapies.

Gene fusions are attractive therapeutic targets (33). Detection of fusions from FFPE is potentially challenging due to low coverage and false-positive calls (2,4,34,35). Interestingly, the three capture methods identified all clinically relevant fusions detected by FF-TruSeq except only one fusion that was not captured by TWIST. *RET* fusions were clinically actionable in two patients with CUP and papillary thyroid cancer. In addition, an *NTRK1* fusion was identified in a colon cancer patient. *NTRK1* fusion is a tumor-agnostic marker with the FDA approved indication of treatment with larotrectinib. The exome capture-based methods provide a high throughput tool that can interrogate FFPE material to profile at a wide-scale the fusions that can guide patient care.

Our study opens the door to use FFPE tissues from archival pathology repositories, a standard-of-care practice worldwide. Fixation and embedding tissues in paraffin are necessary steps for clinical operations as it preserves tissue morphology allowing for histomorphological, immunohistochemical and other *in situ* studies. A particular advantage of FFPE tissues is that they are stored for a longer duration allowing analysis of longer-term patient outcomes (1). The availability of robust transcriptomic data will contribute to the development of patient-derived experimental models. Transcriptomic profiling of tumors can identify specific gene fusions of particular interest to be studied further in pre-clinical models like patient-derived tumor organoids. In addition, samples available from large clinical trials usually come from multiple sites that may not have the infrastructure for freezing tissue, or the collected tissue is inadequate. The FFPE capture-based methods emerge as a suitable method to accomplish correlative studies of such multicenter clinical trials.

To the best of our knowledge, our study is the first to provide a comparison of three different FFPE capture-based methods from the same tumor sample. Previous reports attempted to examine the direct comparison of FF and individual FFPE capture methods from the same sample. These studies had a small size (4-9 tumor samples) (4,36–38). Most of them were limited to examining the gene expression (36,38).

NanoString technologies are a different approach to investigate gene expression and also applicable to FFPE samples. Unlike RNA-seq, which captures the expression of tens of thousands of genes, it is restricted to approximately 800 mRNA targets (39). Using a dataset of 39 FFPE melanoma tumor samples, Kwong *et al*. compared RNA-seq to two NanoString gene expression panels (3). They found that genes with low absolute expression showed poor correlation across platforms. This is consistent with our results across the FFPE capture-based methods. Careful interpretation of the expression profile of low abundant genes should be considered.

The molecular subtype classification was limited by the small sample size of urothelial cancer patients. However, we could identify significant differences across the three capture-based methods. Another consideration of our study is following single-institution conditions of processing. Time to fixation and the degree of degradation of FFPE samples might differ significantly between pathology departments at different institutions. Still, the study reflects the real-world experience and will need to be further validated in multicenter prospective studies.

In conclusion, we compared three capture methods to perform transcriptome profiling using FFPE material using a range of different metrics. In specific functional readouts, such as outlier and immune gene expression, all capture-based methods achieved comparable performance. In other areas, namely, multigene-based subtyping and fusions, there were platform-specific differences. Careful considerations of the biological question must be undertaken to select the ideal RNA capture method of FFPE tumor samples. Altogether, these results point to the possibility of using RNA exome capture-based methods as a promising tool with broad clinical applications.

## Data Availability

The raw RNA-seq datasets analyzed during the current study are available from the corresponding author on reasonable request.

## Acknowledgments

BMF was supported by the Department of Defense CDMRP grant CA160212. This work was also supported by a Conquer Cancer Foundation Long Term International Fellowship Award (KSS) and the Englander Institute for Precision Medicine at WCM (OE, AS, BMF).

## Authors’ contributions

Conception and design of the study (OE, AS, AA, BMF), acquisition and analysis of the data (RB, MS, KS), sequencing of samples (AA, DCW, JZX), statistical and bioinformatic analyses (KS, RB, PD), writing the initial draft of the manuscript (KS, BMF), revise and approve the final version of the manuscript (All authors).

## Supplementary Figures

**Supplementary Figure 1.**
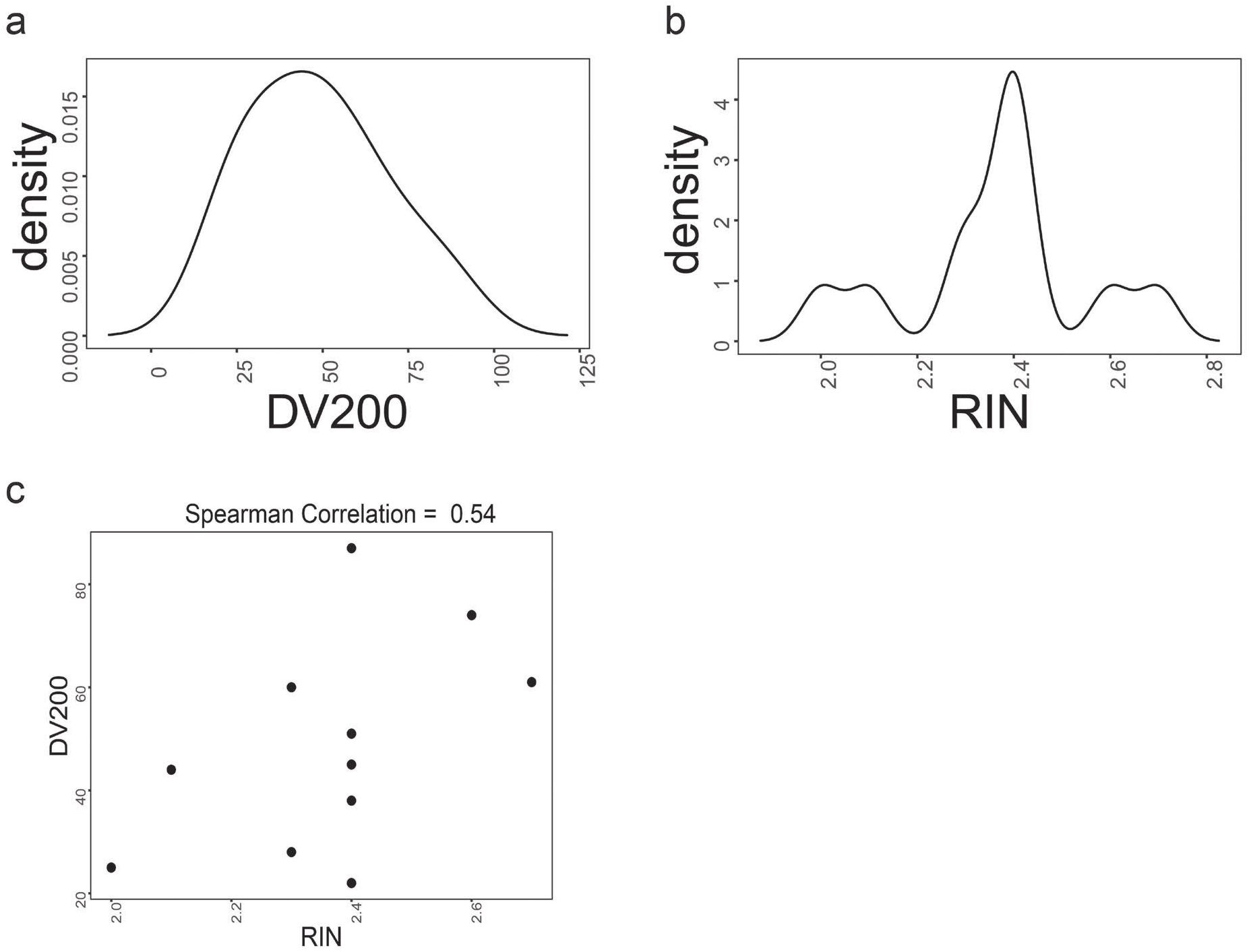
Quality Control Metrics. (a) Density plot of DV200. (b) Density plot of RNA integrity number (RIN). (c) Correlation between DV200 and RIN (Spearman’s correlation = 0.54).

**Supplementary Figure 2.**
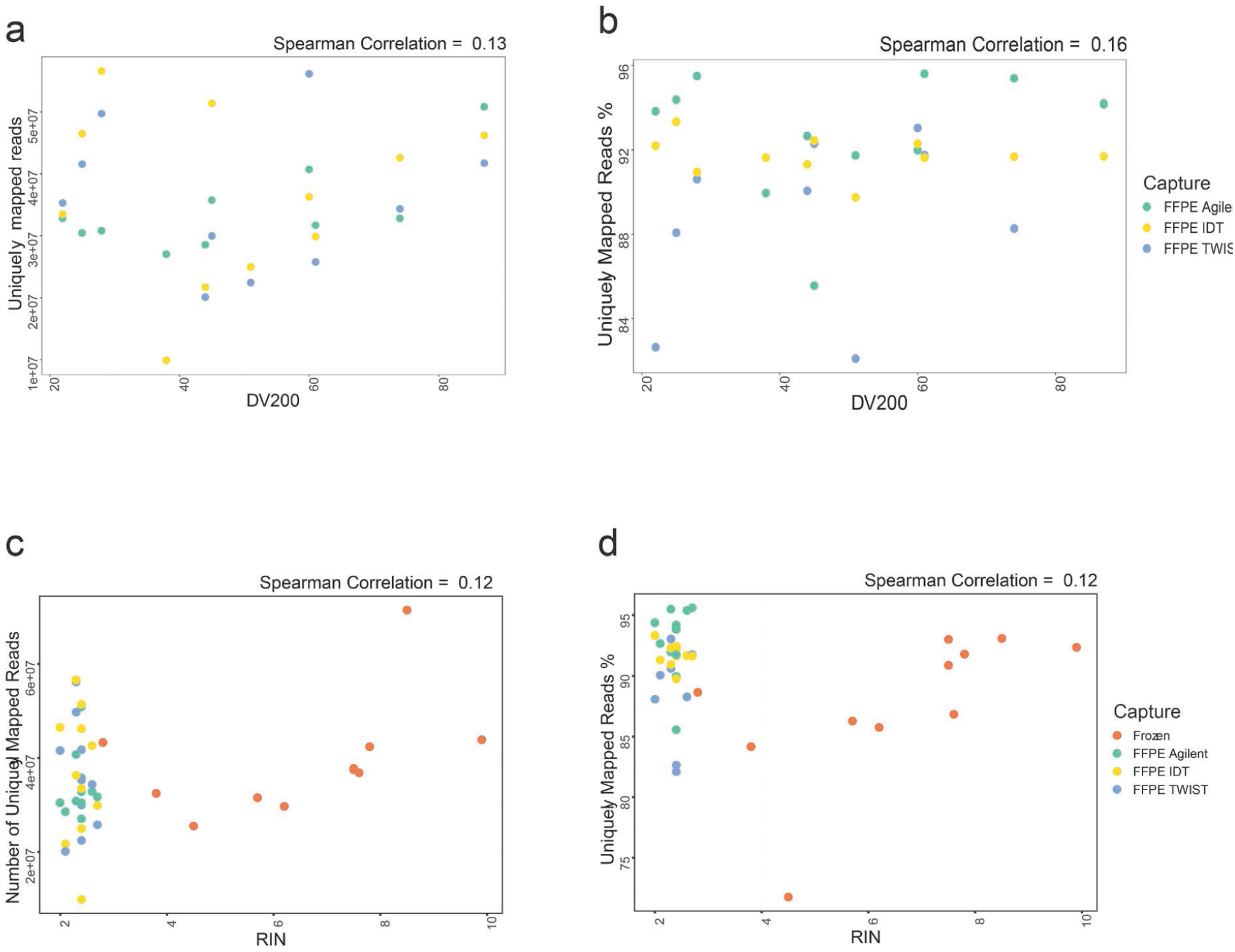
Alignment statistics and correlation with RNA quality control metrics. (a) Correlation between DV200 and uniquely mapped reads (Spearman’s Correlation = 0.13, *p* = 0.51). (b) Correlation between DV200 and Uniquely Mapped Reads % (Spearman’s Correlation = 0.16, *p* = 0.37). (c) Correlation between RIN and Uniquely Mapped Reads (Spearman’s Correlation = 0.12, *p* = 0.45). (d) Correlation between RIN and uniquely mapped reads percentage (Spearman’s Correlation = 0.12, *p* = 0.22).

**Supplementary Figure 3.**
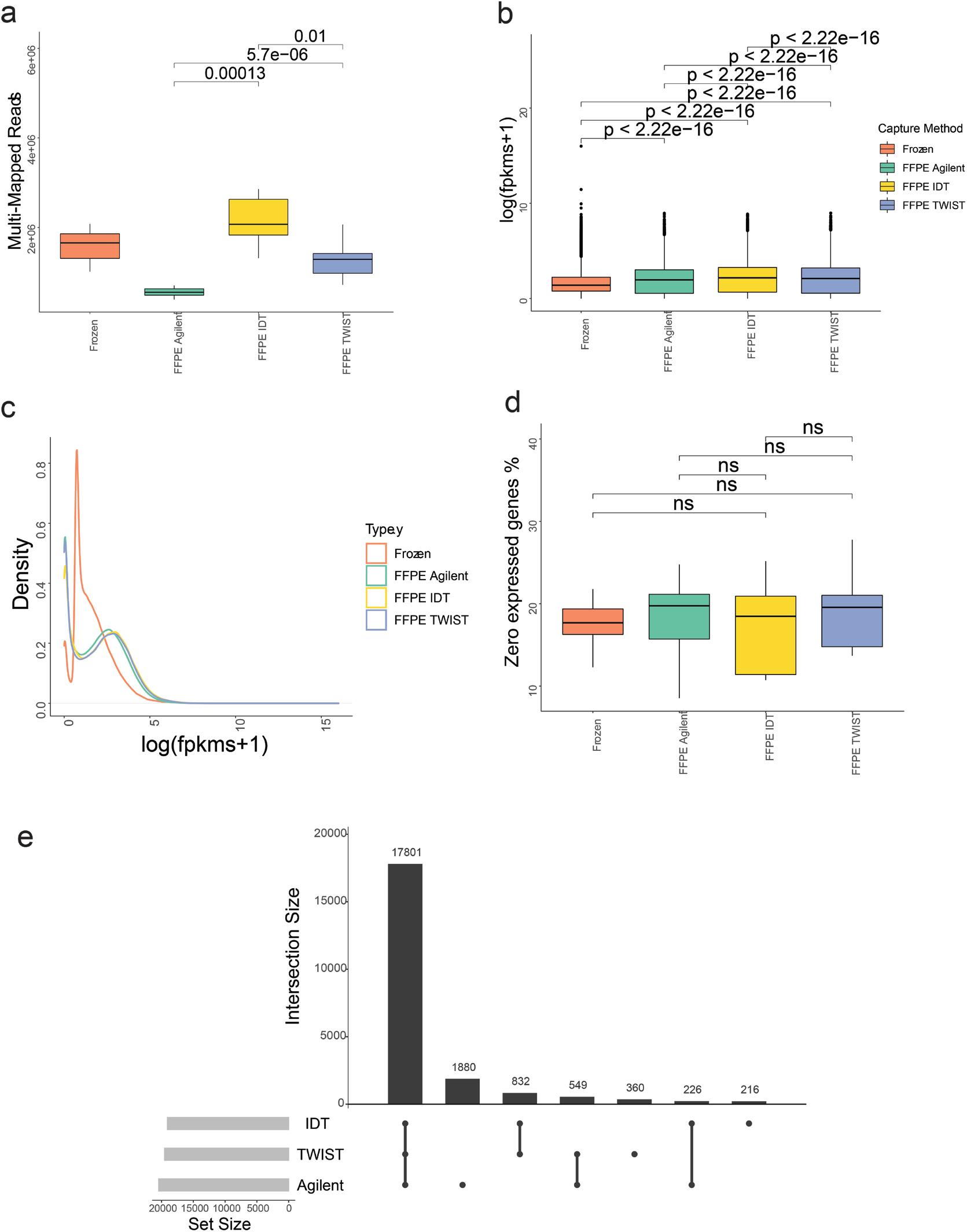
(a) Boxplots of multi-mapping reads comparing the FF-TruSeq and the exome capture methods (Wilcoxon test). (b) Boxplot of log (FPKMS + 1) comparing frozen and the capture methods (Wilcoxon test). (c) Density plot of log (FPKMS +1) comparing frozen and the capture methods. (d) Boxplots of the number of zero expressed genes comparing the frozen and the exome capture methods (Wilcoxon test). (e) Upset plot of the number of genes in the bed file of the capture-based methods showing the overlapping number of captured genes.

**Supplementary Figure 4.**
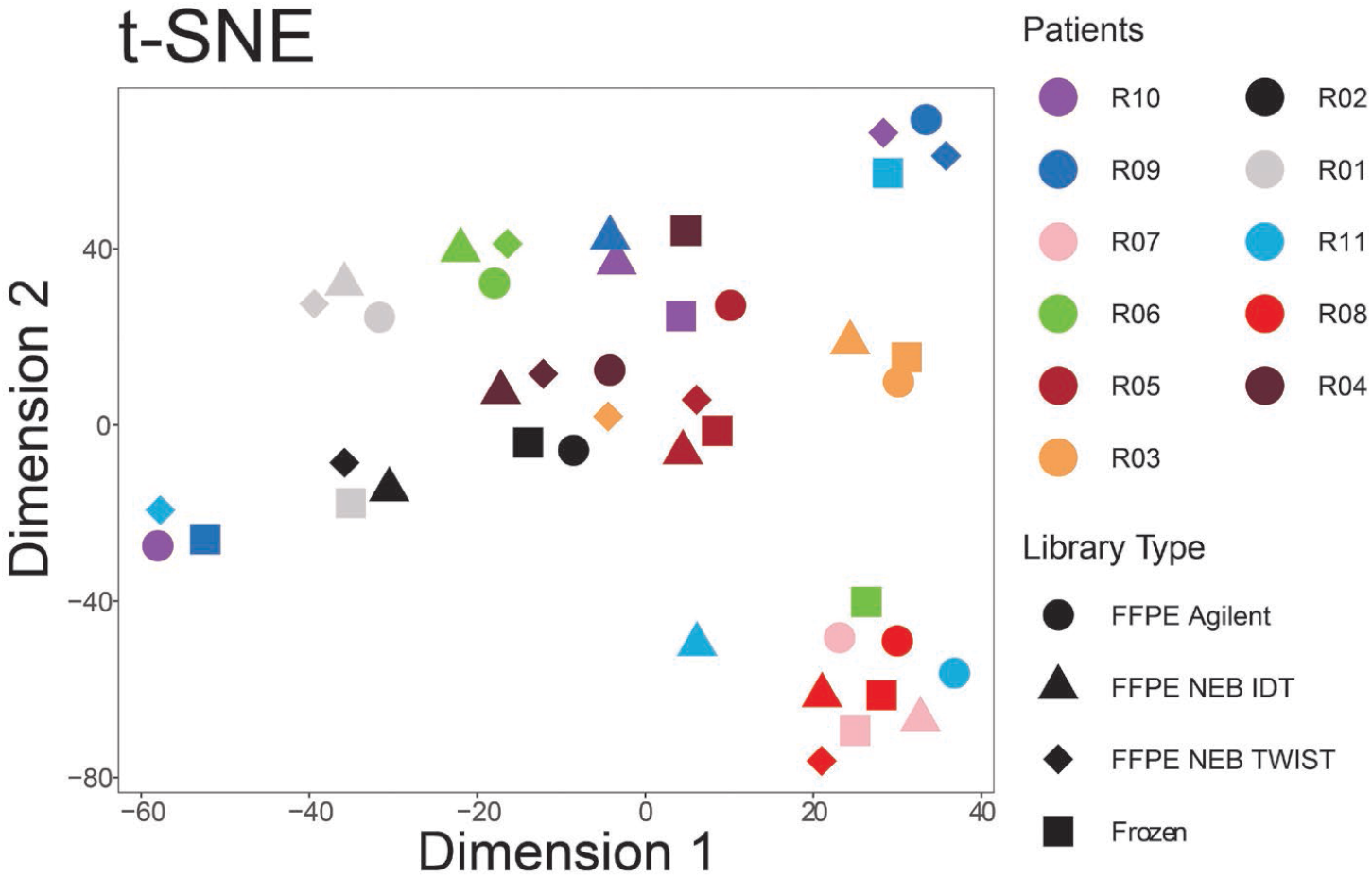
t-SNE projection of samples based on whole transcriptomic profiles (all coding protein-coding genes) colored by patients (Shapes represent different capture methods) showing clustering of FFPE capture-based methods with the FF-TruSeq matched sample.

**Supplementary Figure 5.**
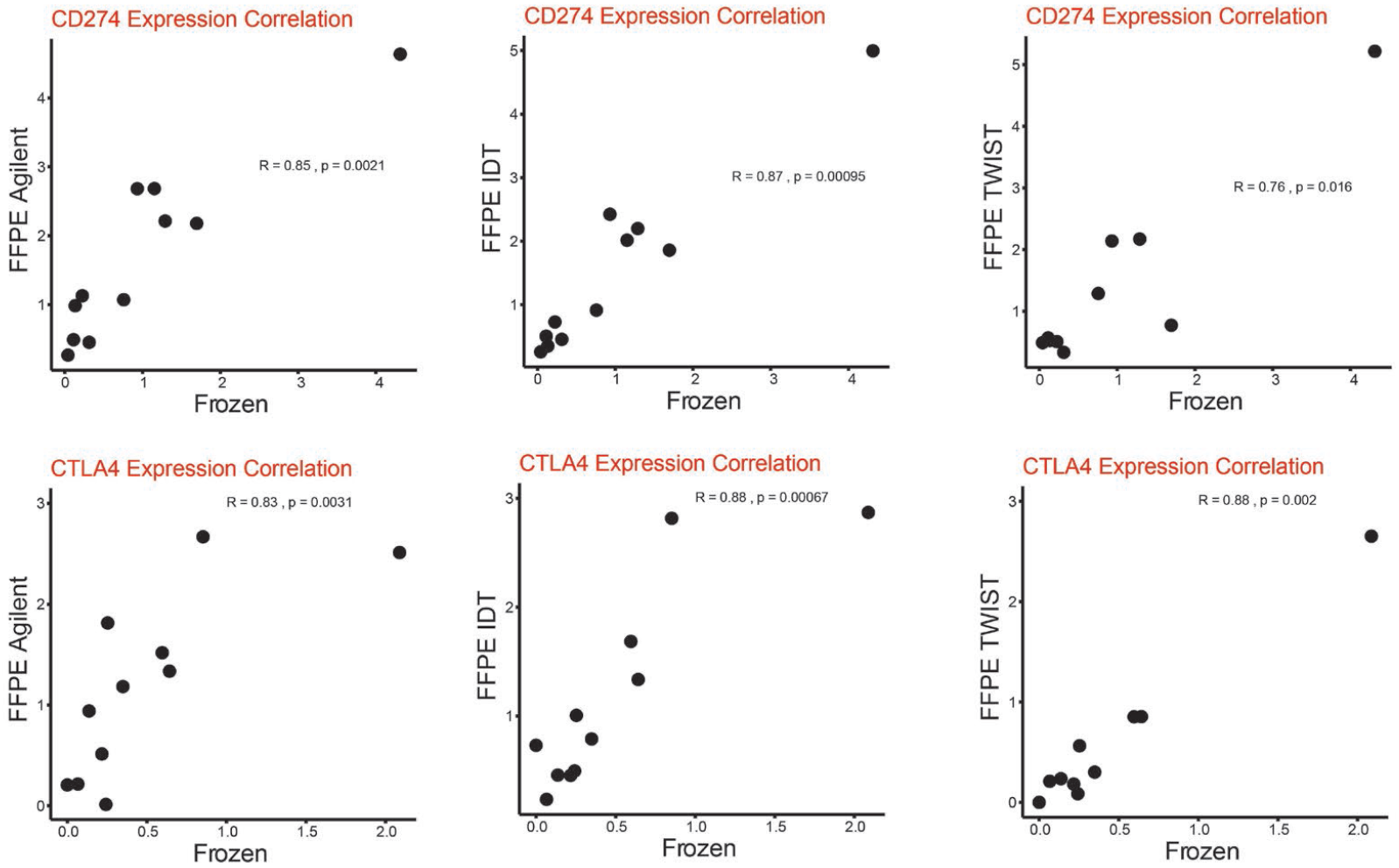
A significant correlation of the FPKM expression of CD274 (PD-L1) and CTLA4 was observed between FF-TruSeq, and the matched three capture-based methods.

## Additional Files

**Supplementary Table 1**: Workflow of RNA sequencing for the fresh-frozen and FFPE tumor samples.

**Supplementary Table 2**: Quality metrics of RNA library preparation from fresh-frozen and FFPE samples

**Supplementary Table 3**: Clinical and pathologic characteristics of patients and their individual samples

**Supplementary Table 4**: Alignment statistics of FFPE capture-based methods and the matched FF-TruSeq samples.

**Supplementary Table 5:** Agreement between consensus molecular classifier using RNA-seq data from FF-TruSeq and matched FFPE capture-based methods from the bladder cancer samples.

**Supplementary Table 6**: Junction read count of the identified gene fusions in FF-TruSeq tumor samples and their matched FFPE exome capture-based method.

